# Estimating the end of the first wave of epidemic for COVID-19 outbreak in mainland China

**DOI:** 10.1101/2020.04.14.20064824

**Authors:** Quentin Griette, Zhihua Liu, Pierre Magal

**Affiliations:** Univ. Bordeaux, IMB, UMR 5251, F-33400 Talence, France., CNRS, IMB, UMR 5251, F-33400 Talence, France; School of Mathematical Sciences, Beijing Normal University, Beijing 100875, People’s Republic of China

**Keywords:** COVID-19 epidemic in mainland China, end of epidemic, reported and unreported cases, control measures

## Abstract

Our main aim is to estimate the end of the first wave epidemic of COVID-19 outbreak in mainland China. We developed mathematical models to predict reasonable bounds on the date of end of the COVID-19 epidemics in mainland China with strong quarantine and testing measures for a sufficiently long time. We used reported data in China from January 20, 2020 to April 9, 2020. We firstly used a deterministic approach to obtain a formula to compute the probability distribution of the extinction date by combining the models and continuous-time Markov processes. Then we present the individual based model (IMB) simulations to compare the result by deterministic approach and show the absolute difference between the estimated cumulative probability distribution computed by simulations and formula. We provide the predictions of the end of the first wave epidemic for different fractions *f* of asymptomatic infectious that become reported symptomatic infectious.

## 1 Introduction

During the outbreak of COVID-19 in China, the government imposed strong intervention measures such as enhanced epidemiological surveys and surveillance, contact tracing, isolation, quarantine. COVID-19 was brought under control in mainland China with these strong measures. Since March 12, the number of daily reported cases imported from mainland China has been kept within 5 for several weeks in mainland China. One of the most concerned issues now is the duration of the epidemic of COVID-19 in mainland China. However, there are several challenges to such analysis. COVID-19 can be contagious during the incubation period. The fraction of asymptomatic infectious cases and unreported cases (with mild symptom) and their contagiousness are of major importance in understanding the evolution of COVID-19 epidemic, and involves great difficulty in their estimation. We refer to Thompson et al. [19] an early article on this topic.

As coronavirus outbreaks surge worldwide, more and more facts [15] show that many new patients which are asymptomatic or have only mild symptoms can transmit the virus. Researches both in [16] and [5] have confirmed that asymptomatic transmission occurs. It has been shown in [21] that some new crown pneumonia patients had higher viral levels in the throat swabs during the early stage of the disease. [14] reported that 13 evacuees from Wuhan, China on chartered flights were infected, of whom 4, never developed symptoms and the estimated asymptomatic proportion in [12] is at 17.9%. A team in China [20] suggests that by February 18, there were 37,400 people with the virus in Wuhan whom authorities didn’t know about. Research in [7] estimates 86% of all infections were undocumented (95% CI: [82%- 90%]) prior to January 23, 2020 travel restrictions. The transmission rate of undocumented infections was 55% of documented infections ([46%-62%]). Due to their greater numbers, undocumented infections were the infection source for 79% of documented cases. The asymptomatic and mild symptomatic cases were missed because authorities aren’t doing enough testing, or ‘preclinical cases’ in which people are incubating the virus but would not be ill enough to seek medical help, would probably slip past screening methods such as temperature checks. The asymptomatic and unreported cases are just going to be really critical for explaining the rapid geographic spread of COVID-19 and indicate containment of this virus will be particularly challenging.

In our previous works on COVID-19 [8], we propose a method applied to the Chinese data to fit the model at the early stage of the epidemic when the number of cases is exponentially growing. In [9, 11] we consider the second phase of the epidemic. Namely, the slowing down of the transmissions. In [10], we estimate the average length of exposure which turns to be very short (6*−* 12 hours). So here we neglect the exposed period. In [4], we consider the model with a discrete age structure by using the data from Japan.

This epidemic model for COVID-19 permits to predict forward in time the future number of cases from early reported case data in regions throughout the world. Here we consider the last phase of first epidemic wave and we evaluated the time of the end of this first wave. Our model incorporates the key features of this epidemic: (1) the importance of the timing and magnitude of the implementation of major government public restrictions designed to mitigate the severity of the epidemic; (2) the importance of asymptomatic infectious, reported (with sever symptom) and unreported (with mild symptom) cases in interpreting the number of reported cases.

This article is devoted to the duration of the epidemic of COVID-19 in mainland China. The duration of the stochastic epidemic has been considered in the 70th by Barbour [2]. We refer to Nishura, Miyamatsu and Mizumoto [13], Lee and Nishiura [6], Thompson, Morgan and Jalava [18] and Britton and Pardoux [3] for more results about stochastic epidemic models. Our goal in the present paper is to investigate the duration of the epidemic of COVID-19 in mainland China in function of the fraction of unreported cases. In reality the epidemic is still present at a low level in China. So, in this article we investigate the extinction time of the disease as long as the model is valid.

## 2 Method

### 2.1 Data

We use the cumulative data of the reported cases confirmed by testing in mainland China from January 20, 2020 to March 18, 2020, taken from the National Health Commission of the People’s Republic of China and Chinese center for disease control and prevention [22, 23]. We should note the following fact: Before February 11, the cumulative data of the reported cases was confirmed by testing. From February 11, the cumulative data included cases that were not tested for the virus, but were clinically diagnosed based on medical imaging. The cumulative data from February 10 to February 15 specified both types of reported cases. But from February 16, the data did not separate the two types of reporting, but reported the sum of both types which makes it impossible for us to know the number of cases tested. There were total 17,409 clinically diagnosed cases from February 10 to February 15. We subtracted 17,409 cases from the cumulative reported cases after February 15 to obtain the approximate data by testing only after February 15 as shown in Table 1 with this adjustment. Note that on January 23^*rd*^ 2020 mainland China started the lock-down of Wuhan city, and implemented other interventions soon on other Chinese cities.

**Table 1:** Cumulative data of reported cases confirmed by testing from January 20, 2020 to March 18, 2020, reported for mainland China.

|  |  |  |  |  |  |  |
| --- | --- | --- | --- | --- | --- | --- |
| **January** |  |  |  |  |  |  |
| 19 | 20 | 21 | 22 | 23 | 24 | 25 |
| 198 | 291 | 440 | 571 | 830 | 1287 | 1975 |
| 26 | 27 | 28 | 29 | 30 | 31 |  |
| 2744 | 4515 | 5974 | 7711 | 9692 | 11791 |  |
| **February** |  |  |  |  |  |  |
| 1 | 2 | 3 | 4 | 5 | 6 | 7 |
| 14380 | 17205 | 20438 | 24324 | 28018 | 31161 | 34546 |
| 8 | 9 | 10 | 11 | 12 | 13 | 14 |
| 37198 | 40171 | 42638 | 44653 | 46472 | 48467 | 49970 |
| 15 | 16 | 17 | 18 | 19 | 20 | 21 |
| 51091 | 70548 – 17409 | 72436 – 17409 | 74185 – 17409 | 75002 – 17409 | 75891 – 17409 | 76288 – 17409 |
| 22 | 23 | 24 | 25 | 26 | 27 | 28 |
| 76936 – 17409 | 77150 – 17409 | 77658 – 17409 | 78064 – 17409 | 78497 – 17409 | 78824 – 17409 | 79251 – 17409 |
| 29 |  |  |  |  |  |  |
| 79824 – 17409 |  |  |  |  |  |  |
| **March** |  |  |  |  |  |  |
| 1 | 2 | 3 | 4 | 5 | 6 | 7 |
| 79824 – 17409 | 79824 – 17409 | 79824 – 17409 | 80409 – 17409 | 80552 – 17409 | 80651 – 17409 | 80695 – 17409 |
| 8 | 9 | 10 | 11 | 12 | 13 | 14 |
| 80735 – 17409 | 80754 – 17409 | 80778 – 17409 | 80793 – 17409 | 80813 – 17409 | 80824 – 17409 | 80844 – 17409 |
| 15 | 16 | 17 | 18 |  |  |  |
| 80860 – 17409 | 80881 – 17409 | 80894 – 17409 | 80928 – 17409 |  |  |  |

### 2.2 The model

The model consists of the following system of ordinary differential equations:

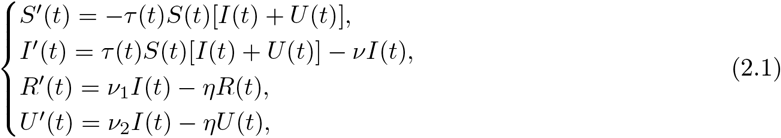

with initial data

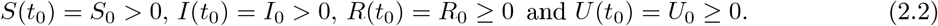

Here *t≥ t*_0_ is time in days, *t*_0_ is the beginning date of the model of the epidemic, *S*(*t*) is the number of individuals susceptible to infection at time *t, I*(*t*) is the number of asymptomatic infectious individuals at time *t, R*(*t*) is the number of reported symptomatic infectious individuals at time *t*, and *U* (*t*) is the number of unreported symptomatic infectious individuals at time *t*. The parameters and initial conditions of the model are given in Table 2 and a flow diagram of the model is given in Figure 1.

**Table 2:**
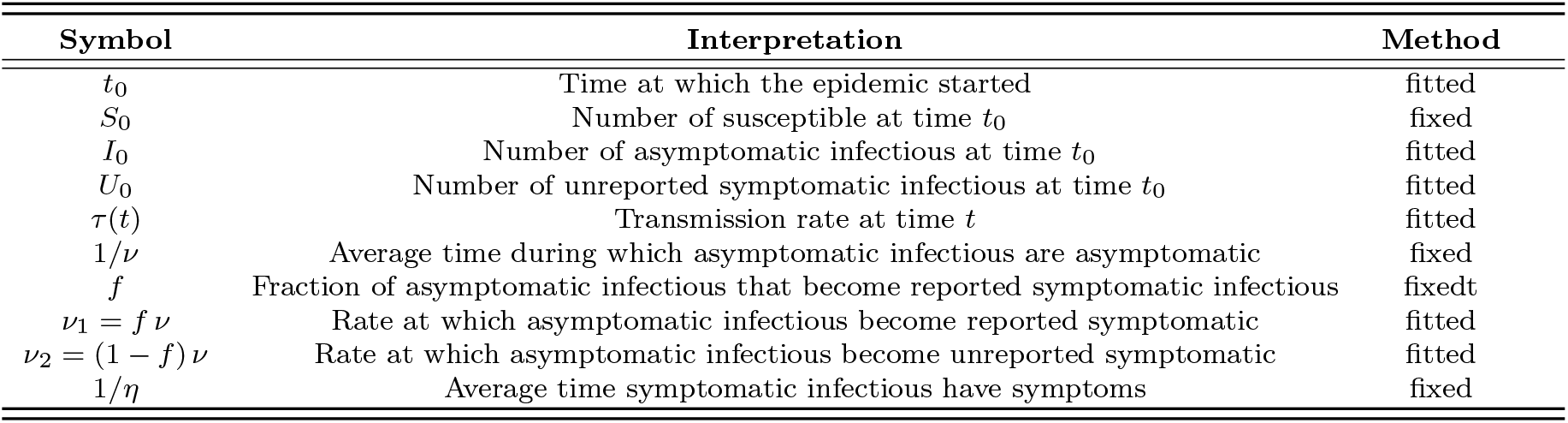
Parameters and initial conditions of the model.

**Figure 1:**
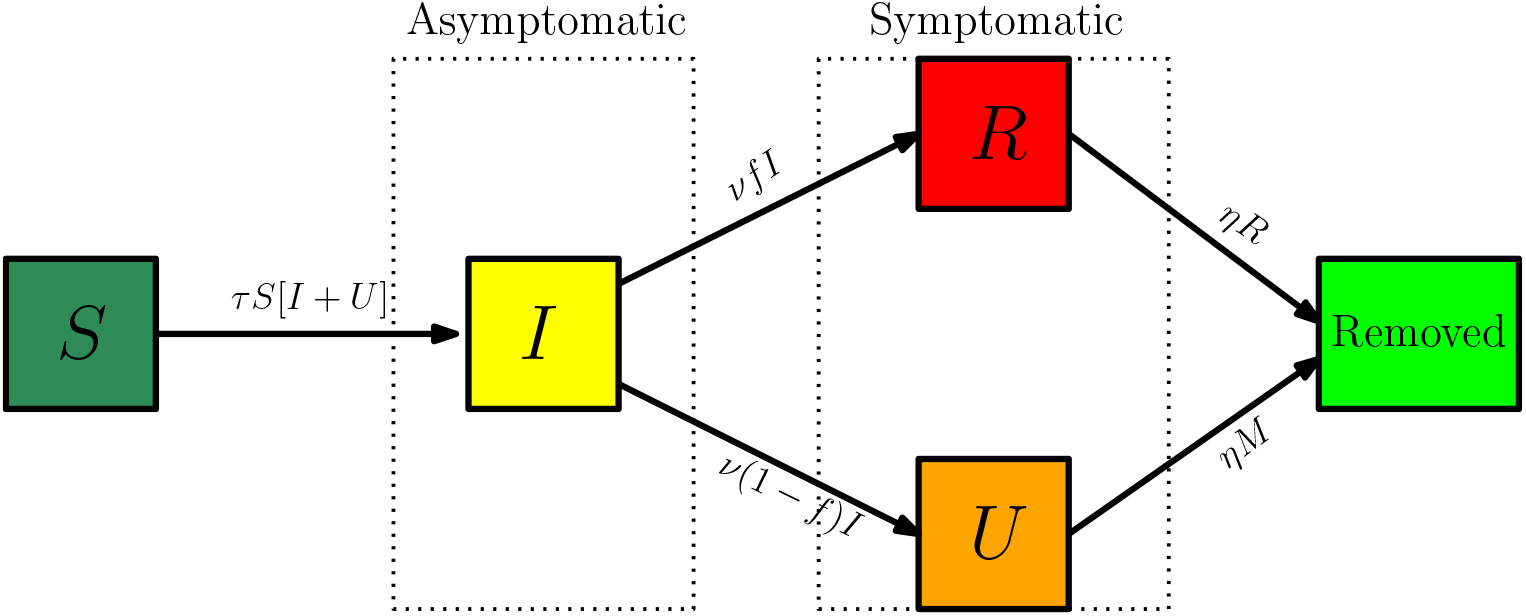
Compartments and flow chart of the model (2.1).

The transmission rate at time *t* is *τ* (*t*). Asymptomatic infectious individuals *I*(*t*) are infectious for an average period of 1*/ν* days. Reported symptomatic individuals *R*(*t*) are infectious for an average period of 1*/η* days, as are unreported symptomatic individuals *U* (*t*). We assume that reported symptomatic infectious individuals *R*(*t*) are reported and isolated immediately, and cause no further infections. The asymptomatic individuals *I*(*t*) can also be viewed as having a low-level symptomatic state. All infections are acquired from either *I*(*t*) or *U* (*t*) individuals. The fraction *f* of asymptomatic infectious become reported symptomatic infectious, and the fraction 1*− f* become unreported symptomatic infectious. The rate asymptomatic infectious become reported symptomatic is *ν*_1_ = *f ν*, the rate asymptomatic infectious become unreported symptomatic is *ν*_2_ = (1 *−f*) *ν*, where *ν*_1_ + *ν*_2_ = *ν*.

During the exponential growth phase *τ* (*t*)*≡ τ*_0_ is constant. We then use a time-dependent decreasing transmission rate *τ* (*t*) to incorporate the effects of the strong measures taken by the authorities to control the epidemics (confinement, contact tracing, etc…). The formula for *τ* (*t*) is

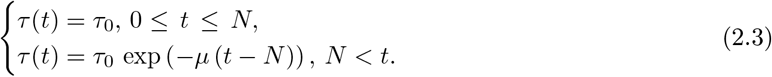

The date *N* and the value of *µ* are chosen so that the cumulative reported cases in the numerical simulation of the epidemic aligns with the cumulative reported case data after day *N*, when the public measures take effect. In this way we are able to project forward the time-path of the epidemic after the government-imposed public restrictions take effect.

The cumulative number of reported cases at time *t* is given by the formula

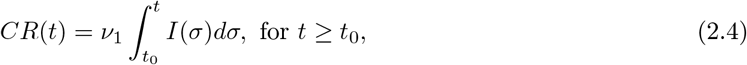

and the cumulative number of unreported at time *t* is given by the formula

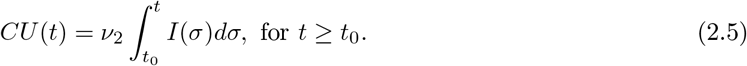

The daily number of reported cases from the model can be obtained by computing the solution of the following equation:

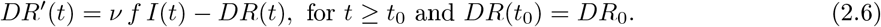

### 2.3 Method to estimate the parameters and initial values of the model

The actual value of *f* is unknown. Because of the strong isolation and testing measures in China, it seems reasonable to take *f* = 0.8 which means that 80% of symptomatic infectious cases go reported. We will however test different values 0.2, 0.4, 0.6, 0.8 of *f*. We assume *η* = 1*/*7, which means that the average period of infectiousness of both unreported symptomatic infectious individuals and reported symptomatic infectious individuals is 7 days. We assume *ν* = 1*/*7, which means that the average period of infectiousness of asymptomatic infectious individuals is 7 days. These values can be modified as further epidemiological information becomes known.

For the exponential growth of reported cumulative cases *CR*(*t*) of the COVID-19 epidemic, we propose a formula:

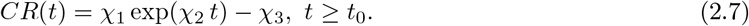

We fix the value of *χ*_3_. The values of *χ*_1_ and *χ*_2_ are fitted to the cumulative reported case data in the exponential growth phase of the epidemic (i.e. we use an exponential fit *χ*_1_ exp(*χ*_2_ *t*) to fit the data *CR*(*t*) + 1). We assume that the initial value *S*_0_ corresponds to the population of the region of the reported case data. The value of the susceptible population *S*(*t*) is assumed to be only slightly changed by the removal of the number of people infected in the beginning of the exponential growth phase. The following formulas for *I*_0_, *U*_0_, *t*_0_, *τ*_0_, and *ℛ*_0_ were derived in [8]. Their numerical values are identified by using (2.8) from the exponential growth phase of the epidemic. The other initial conditions are

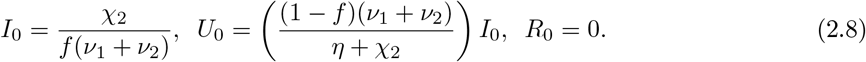

**Remark 2**.**1** *It follows that*

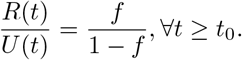

The value of the transmission rate *τ* (*t*), during the exponential growth of the epidemic is the constant value

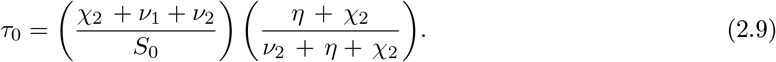

The model starting time of the epidemic is

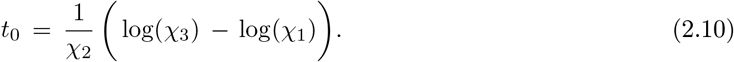

The value of the basic reproductive number is

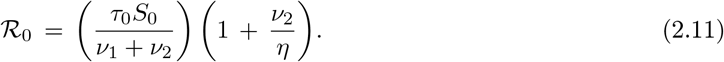

## 3 Result

### 3.1 Derivation of a formula to compute the last day the outbreak

In order to estimate the parameters and initial values of the model, we firstly fix the value *χ*_3_ = 30. The values of *χ*_1_ and *χ*_2_ in *χ*_1_ exp(*χ*_2_ *t*) *− χ*_3_ are fitted to the cumulative reported case data from January 19 to January 26 in Table 1 for mainland China when it is recognized that *CR*(*t*) is growing exponentially. The values of the parameter *τ*_0_ and initial conditions *I*_0_, *U*_0_, *R*_0_,and *t*_0_ are obtained by using formula (2.8)-(2.10). We summarize all the results when *f* takes different values 0.2, 0.4, 0.6, 0.8 in Table 3.

**Table 3:** The parameters χ_1_, χ_2_, χ_3_ are estimated by using the data in Table 1 to fit χ_1_ exp(χ_2_ t)− χ_3_ to the data CR(t) between the following periods January 19 to January 26 for mainland China. The values of I_0_ U_0_, τ_0_, and t_0_ are obtained by using formula (2.8)-(2.10). Here we take χ_3_ = 30 in order to obtain non-zero integer approximation for I_0_, U_0_.

| $\chi_1$ | $\chi_2$ | $\chi_3$ | $t_0$ | $f$ | $\mu$ | $N$ | $I_0$ | $U_0$ | $S_0$ | $\tau_0$ |
| --- | --- | --- | --- | --- | --- | --- | --- | --- | --- | --- |
| 0.2601 | 0.3553 | 30 | 13.3617 | 0.8 | 0.1480 | Jan. 26 | 93.2785 | 5.3494 | $1.40005 \times 10^9$ | $3.3655 \times 10^{-10}$ |
| 0.2601 | 0.3553 | 30 | 13.3617 | 0.6 | 0.1531 | Jan. 26 | 124.3550 | 14.2646 | $1.40005 \times 10^9$ | $3.1920 \times 10^{-10}$ |
| 0.2601 | 0.3553 | 30 | 13.3617 | 0.4 | 0.1574 | Jan. 26 | 186.5325 | 32.0953 | $1.40005 \times 10^9$ | $3.0358 \times 10^{-10}$ |
| 0.2601 | 0.3553 | 30 | 13.3617 | 0.2 | 0.1612 | Jan. 26 | 373.0650 | 85.5875 | $1.40005 \times 10^9$ | $2.8942 \times 10^{-10}$ |

Using the mathematical model (2.1) with parameters and initial values in Table 3, we project the future daily data of reported cases and cumulative data of cases, both reported and unreported for mainland China. In Figures 2 and 3, we present the comparison of the model with the cumulative and daily data for mainland China, respectively.

**Figure 2:**
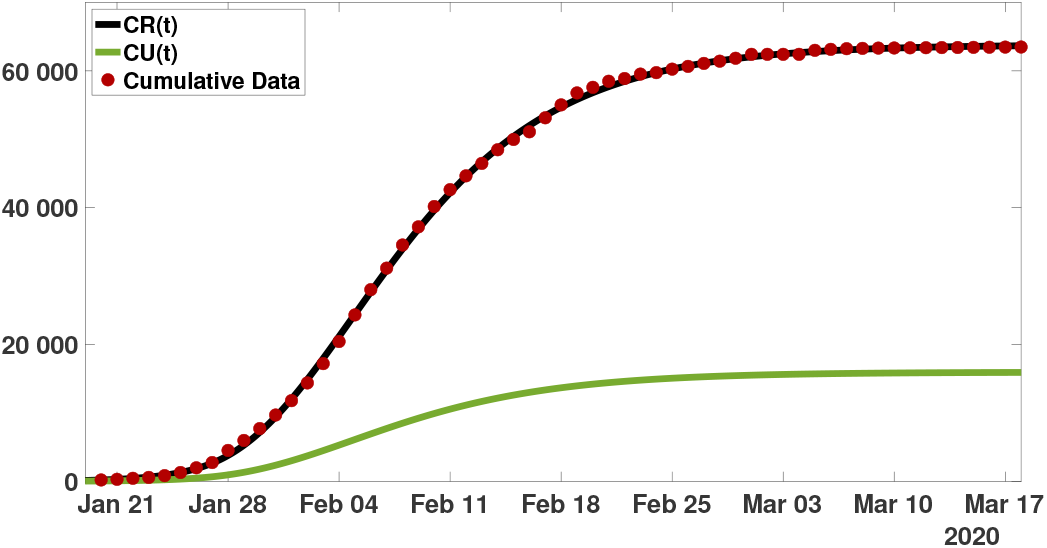
Comparison of the model with the data for mainland China. The parameter values are listed in Table 3 and f = 0.8.

**Figure 3:**
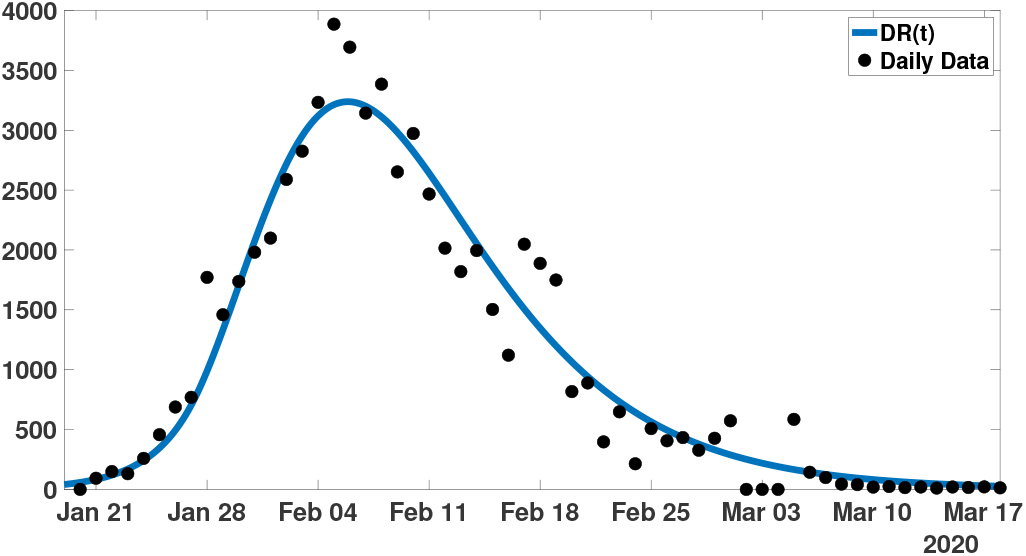
Comparison of the model with the daily data for mainland China. The parameter values are listed in Table 3 and f = 0.8.

The transmission *τ* (*t*) is decreasing exponentially fast for *t > N*. Therefore, if we choose a day *t*_1_ (sufficiently long after the turning point the quantity *τ* (*t*)*S*(*t*) *≤ τ* (*t*)*S*_0_ is small enough) so we can use the approximation

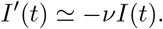

for *I*-equation in system (2.1). This means that the flux of newly infectious can be neglected after the day *t*_1_. We illustrate *S*_0_*τ* (*t*) in Figure 4 for a typical case.

**Figure 4:**
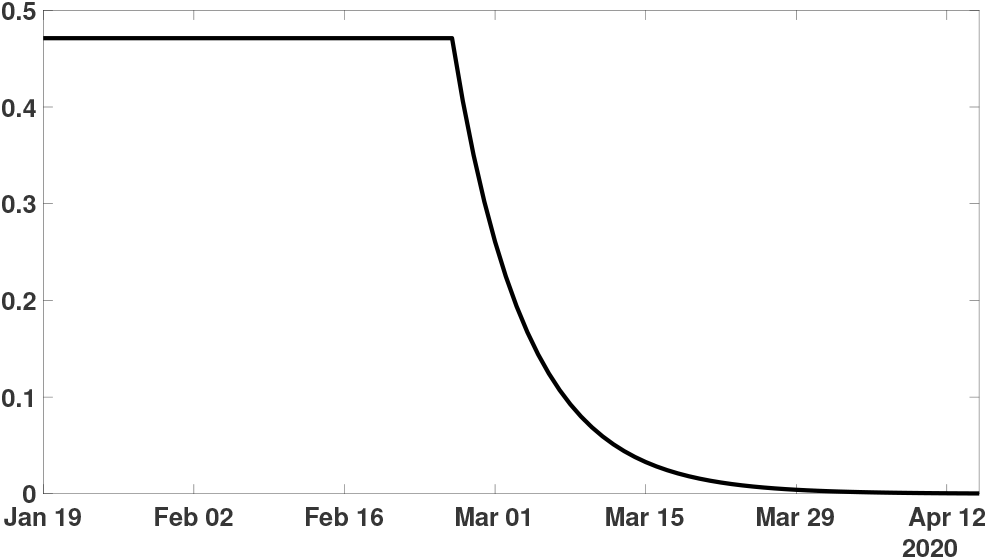
Graph of τ (t)S_0_ = τ_0_S_0_ exp (− µ max (t− N, 0)) with S_0_ = 1.40005 ×10^9^, τ_0_ = 3.3655 ×10^−10^, N = Jan 26, and µ = 0.148. The transmission rate is effectively 0 after March 29. The parameters values correspond the line f = 0.8 in Table 3.

If we assume that this approximation does not influence significantly the number of infectious after the day *t*_1_, we can take *τ* (*t*) = 0 in the original model (2.1) and for *t≥ t*_1_ the resulting system is the following

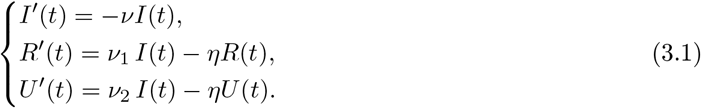

This system is supplemented by the initial data

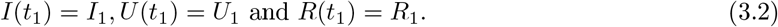

where *I*_1_, *U*_1_ and *R*_1_ are the values of the solutions of the original system (2.1)-(2.2) on day *t*_1_. The flux diagram of model (3.1) is described in Figure 5.

**Figure 5:**
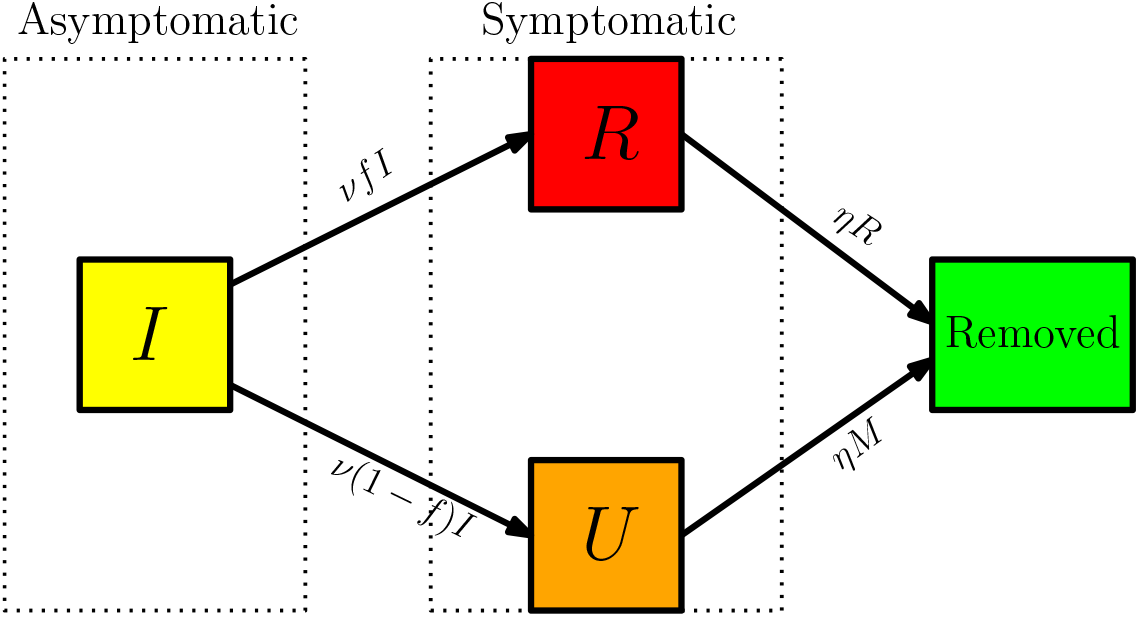
Compartments and flow chart of the model (3.1).

In Figure 6 we represent the error between the solution of (2.1) and the solution of (3.1) for *t > t*_1_ by computing the error as follows.

**Figure 6:**
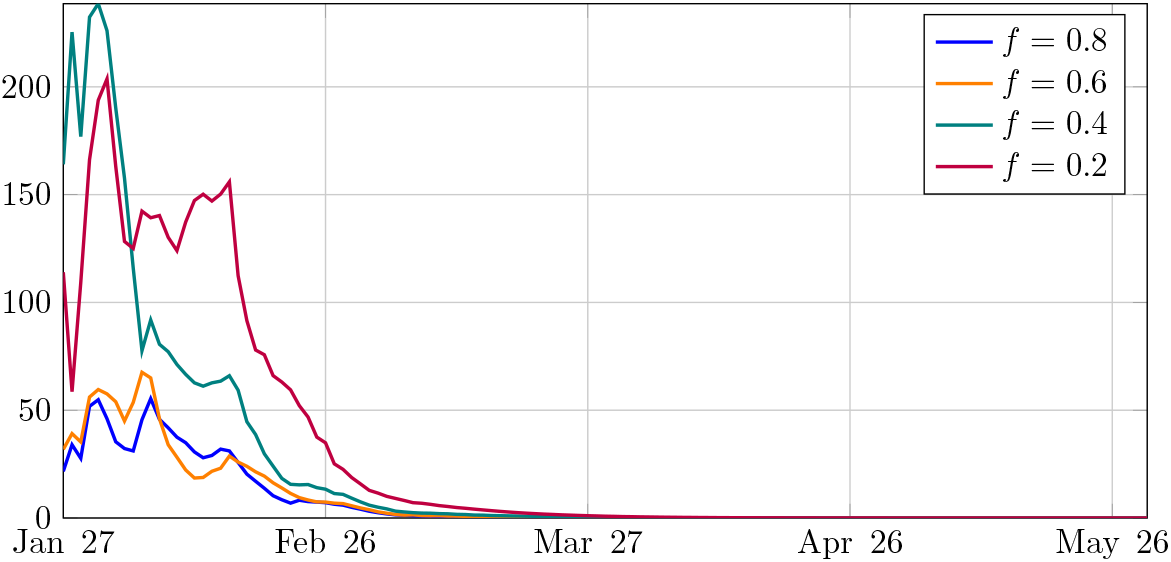
In this figure the x-axis corresponds to t_1_ and the y-axis correponds to the error err(t_1_) defined in (3.3). We observe that the smaller f, the larger the error. Parameter values are listed in Table 3.

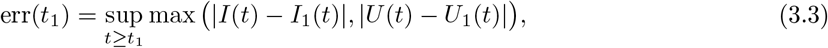

where *I*(*t*) and *U* (*t*) are solution of system (2.1) and *I*_1_(*t*) and *U*_1_(*t*) are solution of system (2.4). This error formula does not involve the component *R*(*t*) for reported cases, because this component is supposed to be known.

In Section 5, we use model (3.1) to compute the probability that no *I*-individual (no asymptomatic infectious) and no *U* -individual (symptomatic unreported) are left after the day *t*. We obtain that there are no more unreported case after the day *t* with the probability

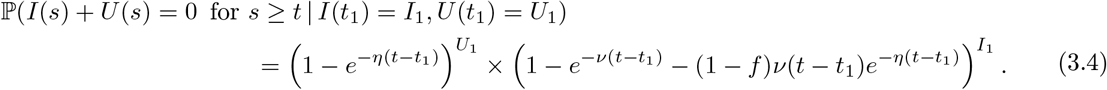

Formula (3.4) allows us to compute the probability of the date of extinction according to the values of *I*(*t*_1_) and *U* (*t*_1_) for different *t*_1_ when *η* = *ν*. (*I*(*t*_1_), *U* (*t*_1_)) is the value of the solution of (2.1) at *t*_1_ with the parameters and initial values taken from Table 3. We show the results in Figure 7. Observe that, as *t*_1_ increases, the probability distribution of the date of extinction seems to converge to a limit profile.

**Figure 7:**
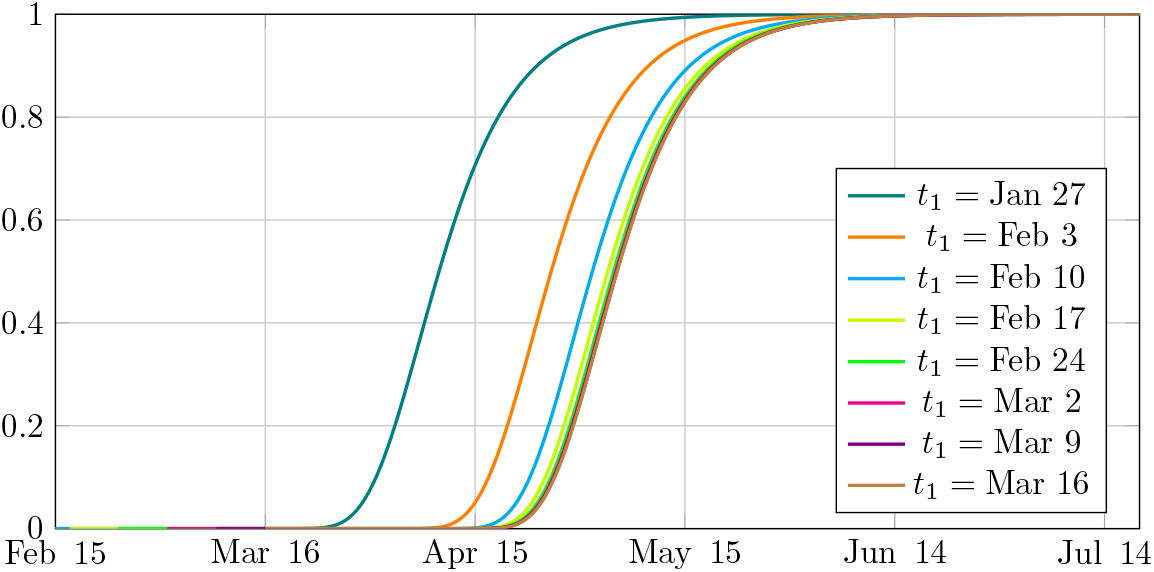
Extinction probability according to formula (3.4). The numerical values for I_1_ and U_1_ were computed from the ODE model at different times, at 7 days intervals since the start of the confinement measures. In this figure we use f = 0.8 and other parameter values are listed in Table 3.

Furthermore, we could also compute 90%, 95% and 99% probability of the date of extinction for different values of *f* by formula (3.4) when *η* = *ν*. In fact, the parameters and initial values in model (2.1) were taken from Table 3 for each value of *f*. Then we compute the values of *I*(*t*_1_) and *U* (*t*_1_) for different values of *t*_1_ which is the value of the solution of (2.1) at *t*_1_. Thus we could compute 90%, 95% and 99% probability of the date of extinction according to the values of *I*(*t*_1_) and *U* (*t*_1_) for different values of *t*_1_ which was summarized in Figure 8.

**Figure 8:**
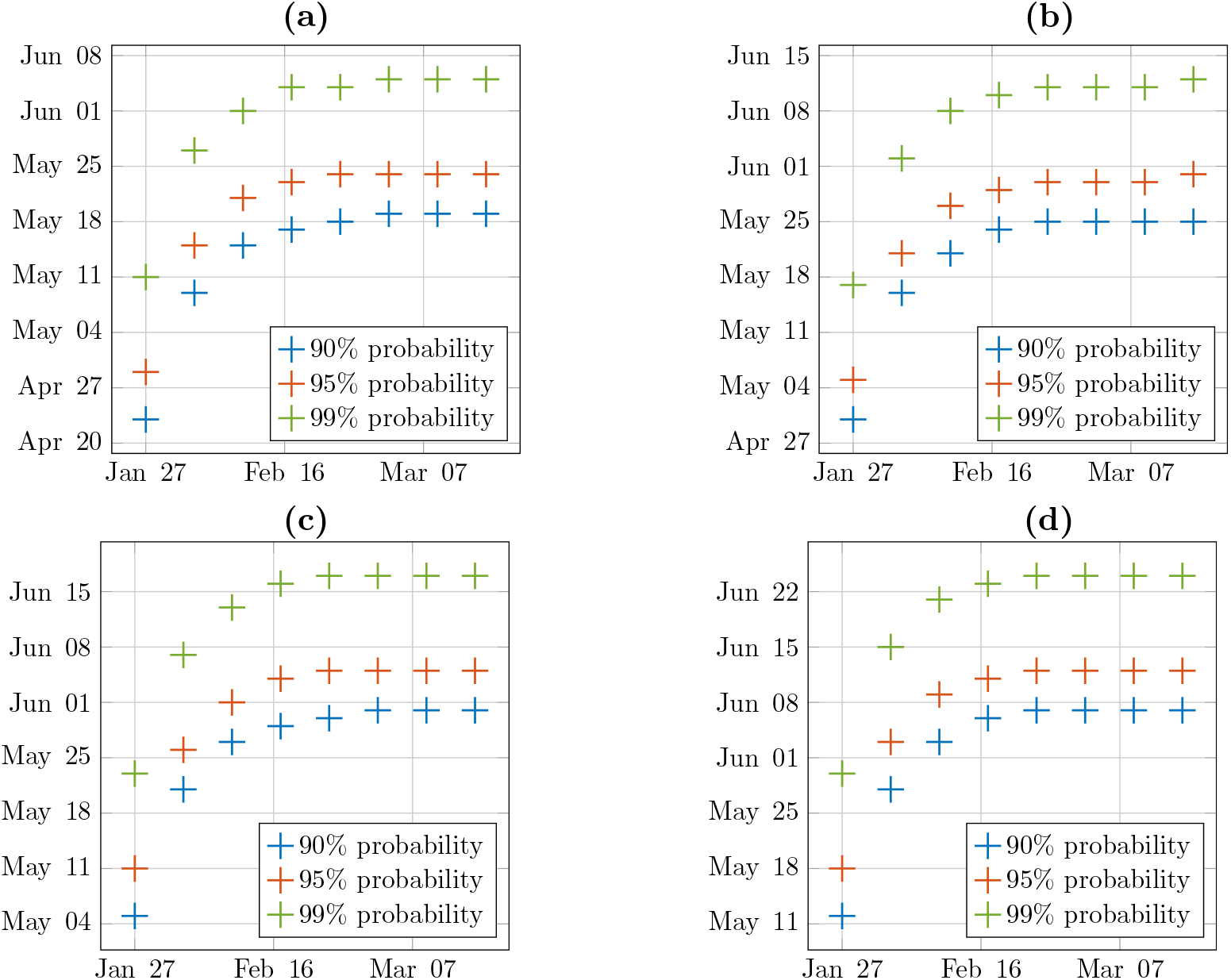
For each figure the x-axis corresponds to the day t_1_ and the y-axis corresponds to the dates of extinction of the disease at different probability level 90%, 95% and 99% computed by using (3.4). We fix f = 0.8 in (a), f = 0.6 in (b), f = 0.4 in (c) and f = 0.2 (d). The values of I_1_ and U_1_ are computed by solving (2.1) up to the time t = t_1_. Parameter values are listed in Table 3.

### 3.2 Stochastic simulations of (2.1) and comparison with (3.4)

To get insight on the variability caused by the randomness of the epidemiological transitions (transmission of the disease due to a contact between an infected and a susceptible, development of symptoms, recovery or death) we developed an individual based model (IMB) in which those epidemiological transitions are modeled by random variables following exponential laws, as described in the flowchart (Figure 5). The interest of these simulations is mostly twofold:

- To estimate the evolution of the epidemic when the accurate number of each class of infected is known. In practice we estimate those numbers by using the deterministic model (2.1) using the available data.
- To give numerical estimates of the cumulative probability distribution of the date of end of the epidemic, without the assumption that *τ* = 0 used in equation (2.1).

In Figure 9, we plot the cumulative distribution for the probability extinction of the epidemic of COVID- 19 obtained by the individual-based simulations. The parameter *t*_1_ in Figure 9 is the date at which the stochastic simulations are started; the precise initial condition is the solution to (2.1) at time *t*_1_. In other words we follow the deterministic model (2.1) up to the date *t*_1_, then start the stochastic simulations.

**Figure 9:**
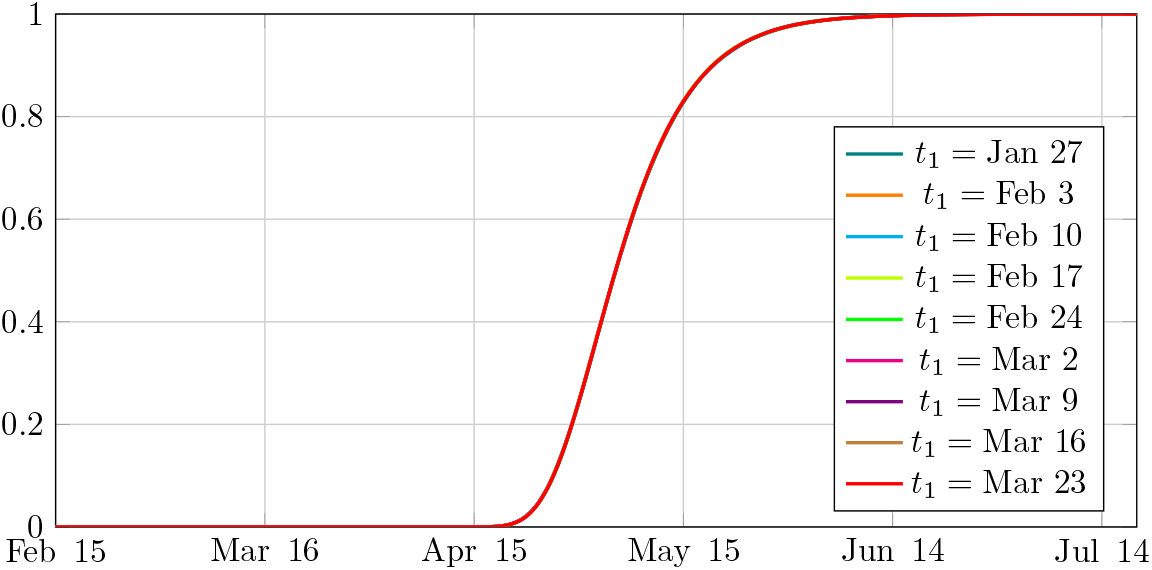
Estimated cumulative probability distributions of the extinction date of the epidemic for different values of the starting point of the stochastic simulations. The red curve is the cumulative distribution corresponding to initial conditions started at t_1_ = 82 (March 23). The initial conditions were computed by rounding the solution to (2.1) at t = t_1_ to the nearest integer. The red curve is estimated with an error of at most 10^−3^ at risk 10^−3^ and other curves are estimated with an error of at most 10^−2^ at a risk of 10^−3^. We took f = 0.8 and other parameter values are shown in Table 3.

The fact that all curves seem to be superimposed with one another indicates that the cumulative probability distribution of the extinction date does not depend on the starting point of the simulations. We also observe that the unique distribution given by the individual-based simulations coincides with the limiting profile for the cumulative distribution in Figure 7. This validates our assumption that *τ* (*t*) can be identified to 0 to compute the cumulative distribution of the extinction date when *t*_1_ is chosen sufficiently large. We infer from Figure 7 that this approximation is acceptable when *t*_1_ is larger than Feb. 17.

To be more precise on the relevance of the approximation formula (3.4), we computed the absolute value of the difference between the cumulative distribution of the extinction date given by (3.4) and the one given by stochastic simulations in Table 4. More precisely, we computed the quantity

**Table 4:** Absolute difference between the cumulative distribution given by the stochastic simulations and the approximation (3.1). For each t_1_ we computed the cumulative distributions with a risk 10^−3^ of an error greater than 10^−2^, starting from an initial condition given at t = t_1_. This corresponds to a total of n = 152019 independent simulations for each set of initial conditions. For each t_1_, the initial condition was computed by rounding the solution to (2.1) at t = t_1_ to the nearest integer.

|  |  |  |  |  |  |  |
| --- | --- | --- | --- | --- | --- | --- |
| $t_1$ | 26 | 33 | 40 | 47 | 54 | 61 |
| date | Jan. 27 | Feb. 3 | Feb. 10 | Feb. 17 | Feb. 24 | Mar. 2 |
| diff($t_1$) | $8.6 \times 10^{-1}$ | $4.4 \times 10^{-1}$ | $1.7 \times 10^{-1}$ | $6.4 \times 10^{-2}$ | $2.5 \times 10^{-2}$ | $8.1 \times 10^{-3}$ |
| $t_1$ | 68 | 75 | 82 | | | |
| date | Mar. 9 | Mar. 16 | Mar. 23 |  |  |  |
| diff($t_1$) | $3.5 \times 10^{-3}$ | $8.5 \times 10^{-4}$ | $5.7 \times 10^{-4}$ | | | |

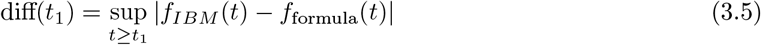

for each *t*_1_ presented in Figures 7 and 9, where *f*_*IBM*_ is the cumulative distribution computed by stochastic simulations (Figure 9) and *f*_formula_ is the cumulative distribution given by (3.4) (Figure 7).

Finally, we compared the results of the individual based model simulations starting from the to the result of the model (2.1). The plots of the average value over our individual-based simulations compared to the corresponding component of the model (2.1) are presented in Figure 10. In Figure 11 we present a representation of the average and standard deviation of the populations computed by the individual-based simulations. Note however that the high variability observed is largely due to the small size of the initial population at *t*_0_. In Table 5 we show that this variability diminishes when the starting time of the stochastic simulations increases.

**Table 5:** Maximal standard deviation for the components I, R and U computed by stochastic simulations started at date t_1_ with initial condition given by the solution to (2.1) with the parameters from Table 3. The ODE model (2.1) is solved up to t = t_1_, and we take the solution to (2.1) at t = t_1_ as initial condition for the stochastic simulations. σ(t) is the maximum, at time t, of the standard deviations of the quantities I(t), R(t) and U (t) in a sample of n = 1000 independent simulations started at t = t_1_, and is expressed in number of individuals. We took f = 0.8 and other parameters are taken from Table 3.

|  |  |  |  |  |  |  |
| --- | --- | --- | --- | --- | --- | --- |
| $t_1$ | $t_0$ | 18 | 22 | 26 | 33 | 40 |
| date | Jan. 14 | Jan. 19 | Jan. 23 | Jan. 27 | Feb. 3 | Feb.10 |
| $\max_{t \geq t_1} \sigma(t)$ | 3717 | 1685 | 787 | 401 | 186 | 106 |

**Figure 10:**
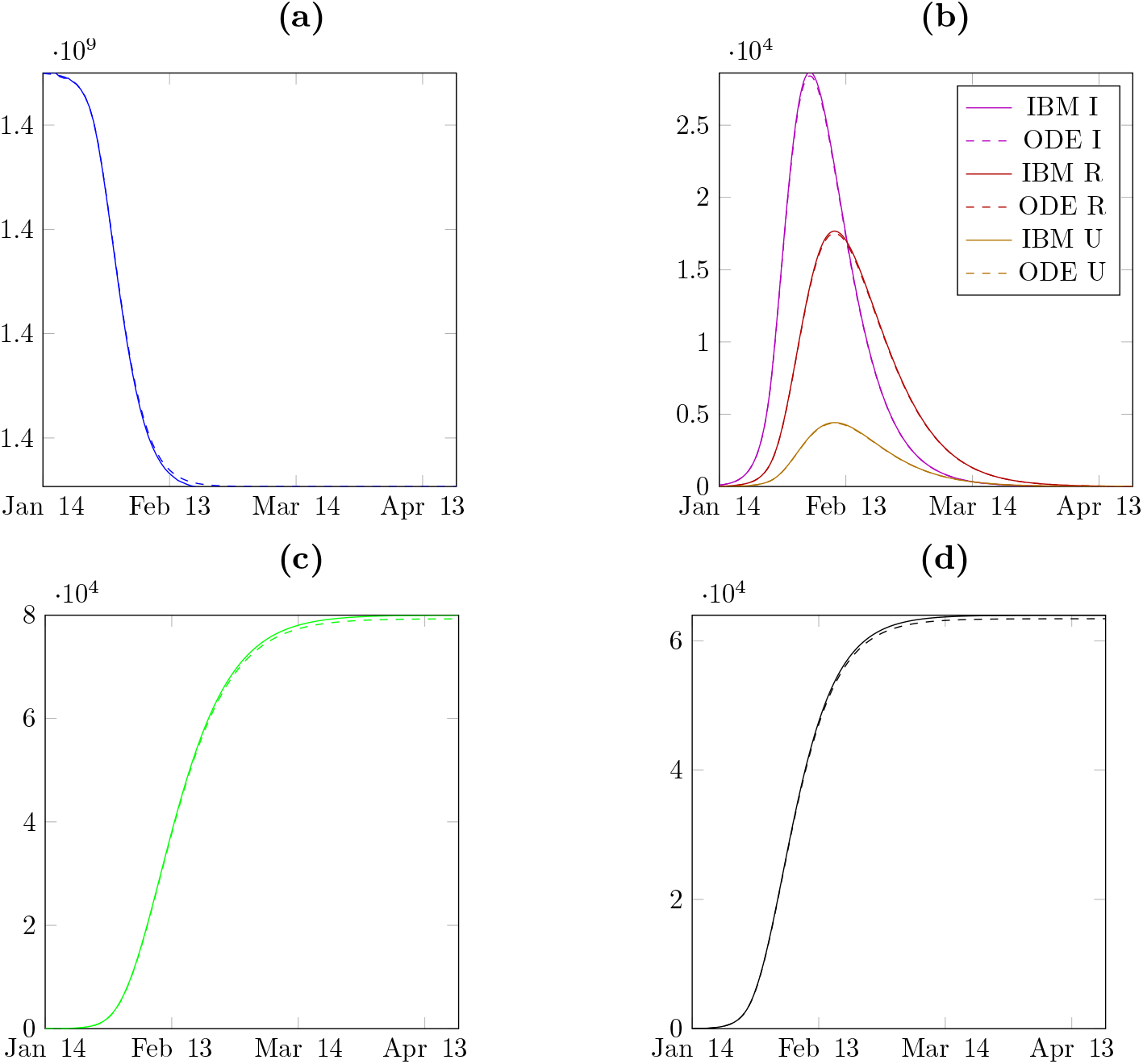
In figure (a) we plot a comparison between the average S (susceptible) computed from the IBM and the S component of the solution of (2.1). In figure (b) we plot a comparison between the average I (asymptomatic), R (reported) and U (unreported) computed from the IBM and the components I, R and U of the solution of (2.1). In figure (c) we plot a comparison between the average RR (removed) computed from the IBM and the components RR of the solution of (2.1). In figure (d) we plot a comparison between the average CR (cumulative reported cases) computed from the IBM and the curve CR computed by (2.1)-(2.4). In this figure 500 independent runs of the IBM simulations are used and the corresponding components of the ODE model start from the same initial condition (at t = t_0_). The parameters we used for both computations are the following: I_0_ = 93, U_0_ = 5, S_0_ = 1.40005 × 10^9^ − (I_0_ + U_0_), R_0_ = RR_0_ = CR_0_ = 0 and f = 0.8, τ_0_ = 3.3655 × 10^−10^, N = 26, µ = 0.148,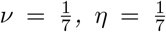, t_0_ = 13.3617.

**Figure 11:**
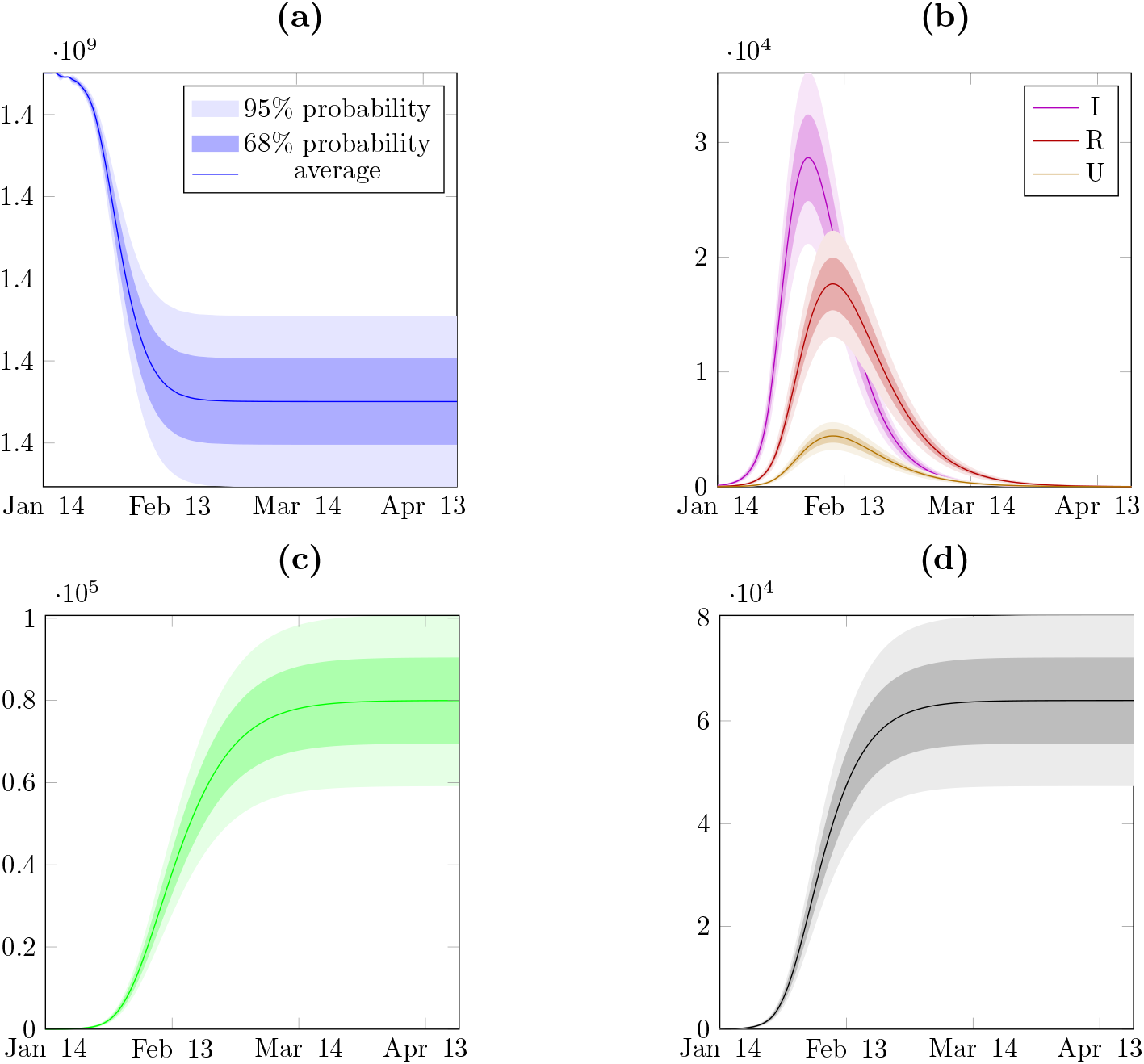
In figure (a) we plot the mean value and variance of S (susceptible) computed from the IBM. The dark blue area contains 68% of the trajectories, and the light blue area 95%. In figure (b) we plot the mean value and variance of I (infected), R (reported) and U (unreported) computed from the IBM. The dark areas contains 68% of the trajectories, and the light areas 95%. In figure (c) we plot the mean value and variance of RR (removed) computed from the IBM. The dark green area contains 68% of the trajectories, and the light green area 95%. In figure (d) we plot the mean value and variance of CR (cumulated reported) computed from the IBM. The dark gray area contains 68% of the trajectories, and the light gray area 95%. We use 500 independent runs of the IBM simulations. The parameters we used for both computations are the following: I_0_ = 93, U_0_ = 5, S_0_ = 1.40005 × 10^9^ − (I_0_ + U_0_), R_0_ = RR_0_ = CR_0_ = 0 and f = 0.8, τ_0_ = 3.3655 × 10^−10^, N = 26, µ = 0.148,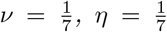, t_0_ = 13.3617.

## 4 Discussion

In this study we mixed the deterministic approach, which correctly describes the initial and intermediate phases of the epidemics, with individual-based models which give estimates on the real extinction date of the epidemics. In Table 6 we summarize our findings for *f* = 0.8, 0.6, 0.4 and 0.2. From this table we deduce that the larger *f* is the earlier the epidemic will stop. Therefore it is very important to increase as much as possible the value of *f* in order to reduce the duration of the epidemic of COVID-19 in mainland China.

**Table 6:** In this table we record the last day of epidemic obtained from Figure 8 by fixing t_1_ to March 16.

| Level of risk | 10% | 5% | 1% |
| --- | --- | --- | --- |
| Extinction date ($f = 0.8$) | May 19 | May 24 | June 5 |
| Extinction date ($f = 0.6$) | May 25 | May 31 | June 12 |
| Extinction date ($f = 0.4$) | May 31 | June 5 | June 17 |
| Extinction date ($f = 0.2$) | June 7 | June 12 | June 24 |

We developed a mathematical framework to predict reasonable bounds on the date of end of the COVID-19 epidemics in mainland China, provided quarantine and confinement measures are maintained with sufficient strength. In particular, the day at which confinement was eased is nowhere near any reasonable bound for the extinction date. Therefore, a secondary outbreak in mainland China is not to be discarded: there is a high probability that there still exists a significant number of unreported infected individuals in the population.

Many parameters are still unknown concerning the future behavior of the pandemics. For what concerns mainland China, even if the remaining hidden number of infected individuals can be estimated by our models, the transmission rate after the end of the confinement measures remains unknown. Indeed, it is reasonable to expect that sociological phenomena like the awareness of the danger have a strong impact on this quantity, because people will tend to avoid risky behavior. There is a strong incentive to identify quantitatively this transmission rate after the end of confinement measures, as we believe that this parameter is crucial to determine whether the epidemic will potentially start again or not. This issue will be addressed in a forthcoming paper.

In this article we computed the end day of the epidemic by neglecting the fact that complete confinement has been progressively lifted very early in the history of the epidemics, with Chinese people going back to work as early as February 10th. Indeed the data from Table 7 and 8 show a number of daily new contaminations occurring inside the territory which is very low since mid-March (less than 10 people a day, Table 8) and the majority of daily new contaminations actually come from abroad (Table 7). This seems to indicate that the propagation inside the country has stopped and the bulk of new contaminations are due to imported cases from abroad. These numbers are relatively surprising compared to our model. In our model, we are quite optimistic since we have placed ourselves in the hypothesis of a very strong confinement, as if the initial shutdown had been respected throughout the epidemics until the very last day. However, we still predict more than 100 new reported cases a day until April 3rd. In Italy and South Korea, by comparison, our predictions stay consistent with the observed data [11].

**Table 7:** Daily data of reported confirmed cases imported from abroad from March 4, 2020 to April 9, 2020.

| March |  |  |  |  |  |  |  |  |  |  |  |  |  |  |  |
| --- | --- | --- | --- | --- | --- | --- | --- | --- | --- | --- | --- | --- | --- | --- | --- |
| 4 | 5 | 6 | 7 | 8 | 9 | 10 | 11 | 12 | 13 | 14 | 15 | 16 | 17 | 18 | 19 |
| 2 | 16 | 24 | 3 | 4 | 2 | 10 | 6 | 3 | 7 | 16 | 12 | 20 | 12 | 34 | 39 |
| 20 | 21 | 22 | 23 | 24 | 25 | 26 | 27 | 28 | 29 | 30 | 31 |  |  |  |  |
| 41 | 45 | 39 | 74 | 47 | 67 | 54 | 54 | 44 | 30 | 48 | 35 |  |  |  |  |
| April |  |  |  |  |  |  |  |  |  |  |  |  |  |  |  |
| 1 | 2 | 3 | 4 | 5 | 6 | 7 | 8 | 9 |  |  |  |  |  |  |  |
| 35 | 29 | 18 | 25 | 38 | 32 | 59 | 61 | 38 |  |  |  |  |  |  |  |

**Table 8:** Daily data of reported confirmed cases reported in mainland China from March 4, 2020 to April 9, 2020.

| March |  |  |  |  |  |  |  |  |  |  |  |  |  |  |  |
| --- | --- | --- | --- | --- | --- | --- | --- | --- | --- | --- | --- | --- | --- | --- | --- |
| 4 | 5 | 6 | 7 | 8 | 9 | 10 | 11 | 12 | 13 | 14 | 15 | 16 | 17 | 18 | 19 |
| 137 | 127 | 75 | 41 | 36 | 17 | 14 | 9 | 5 | 4 | 4 | 4 | 1 | 1 | 0 | 0 |
| 20 | 21 | 22 | 23 | 24 | 25 | 26 | 27 | 28 | 29 | 30 | 31 |  |  |  |  |
| 0 | 1 | 0 | 4 | 0 | 0 | 1 | 0 | 1 | 1 | 0 | 1 |  |  |  |  |

| April |  |  |  |  |  |  |  |  |
| --- | --- | --- | --- | --- | --- | --- | --- | --- |
| 1 | 2 | 3 | 4 | 5 | 6 | 7 | 8 | 9 |
| 0 | 2 | 1 | 5 | 1 | 0 | 3 | 2 | 4 |

Although schools and universities are still closed, the increase in the number of contacts due to workers going back to factories surely increases the transmission rate compared to a total shutdown. Because of this, our estimate of the date of end is very optimistic and the actual date of end should be event later in the future. In the worse case scenario the epidemic may start again. Hopefully some other phenomenon somehow leads to an early end, like the evolution of temperature and humidity with the approach of summer (some influence of those factors in COVID-19 transmission has been remarked in recent works, see *e*.*g*. [17]). In particular dry and hot weather may be favourable to the extinction of the disease.

To conclude the discussion, we should mention a possible alternative approach by using the Kolmogorov equation (see Allen [1] and Britton and Pardoux [3]). This is left for future work.

## 5 Supplementary

### 5.1 Formula to compute the probability distribution of the extinction date

We use continuous-time Markov processes to compute the exact distribution of the date of end of the epidemic after the transmission rate is effectively taken as zero. We start on *t*_1_ with initial values *I*_1_, *U*_1_, and *R*_1_ for *I*-individuals, *U* -individuals and *R*-individuals, respectively. The evolution of each individual is guided by independent exponential processes, and we have the following:

i. Each individual *I* will change state following an exponential clock of rate *ν*. When *I* changes its state, it will be transferred to the class of *R*-individuals with probability *f* and to the class of *U* -individuals with probability (1 *− f*);
ii. Each individual in the state *U* will change state following an exponential clock with rate *η* and become removed individual;
iii. Each individual in the state *R* will change state following an exponential clock with rate *η* and become removed individual

Since the class *I* has only outgoing fluxes, the law of extinction for the *I*-individuals is

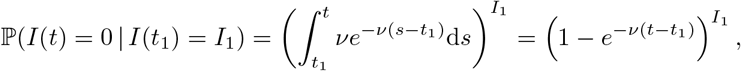

and the probability to have some *I*-individual left at time *t* is

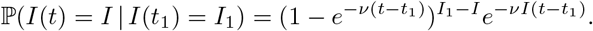

For the *U* -individuals and the *R*-individuals, the situation is more intricate. Indeed, the *U* -individuals and the *R*-individuals vanish at a constant rate *η* but new individuals appear from the *I* class at rate (1*− f*)*ν* and *fν*, respectively, depending on the remaining stock of *I*. Therefore the probability that *U* gets extinct before *t* also depends on the number of remaining *I*. It is actually easier to compute directly the extinction property for the sum *I* + *U*, which is our aim anyways.

When *ν ≠ η*, we obtain

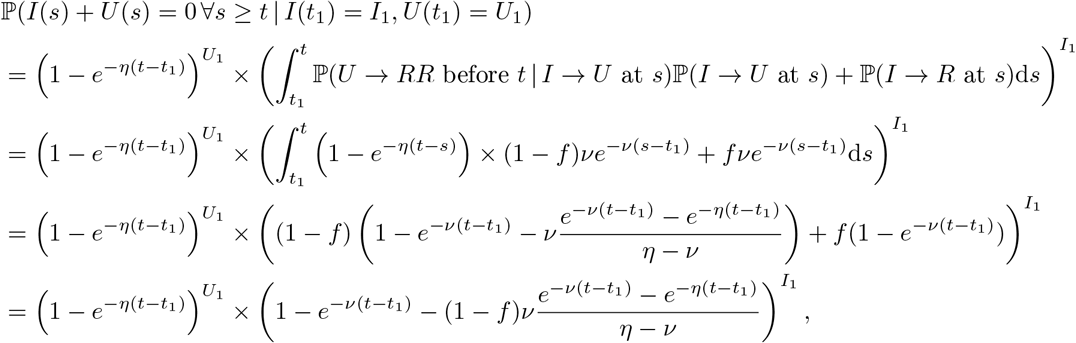

where the *RR*-individuals are the removed individuals.

Similarly when *η* = *ν*, we obtain

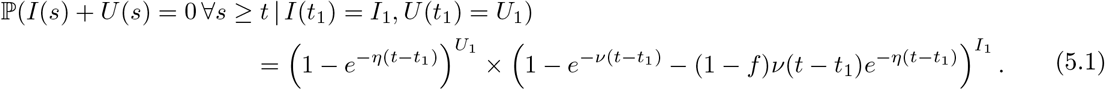

### 5.2 Cumulative distribution of the date of end of the epidemic

The stochastic simulations introduced in section 3.2 can be used, in particular, to precisely estimate the cumulative probability distribution of the date of end of the epidemic, defined as the last time at which the quantity *I* + *U* is positive.

In order to get a measure of the precision we remark that the values taken by the cumulative probability distribution *f* (*t*) can be estimated by the average of independent measures of the random variable

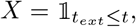

which follows an Bernouilli distribution of parameter *f* (*t*). Consecutive runs of the individual-based simulations yield independent observations *X*_*n*_ of this distribution. By Hoeffding’s inequality we have for all *ε >* 0 and *n ∈* N

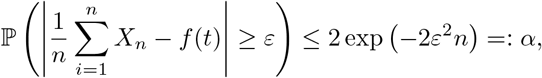

and we achieved an error of at most *ε* = 10^*−*3^ at risk *α ≤* 10^*−*3^ by running 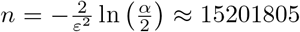 independent individual-based simulations to estimate the probability distribution of the extinction time (Figure 9, *t*_1_ = 82 *i*.*e*. March 23). Other curves are esimated on the basis of 152019 independent simulations, which amouts to an error of at most 10^*−*2^ at risk 10^*−*3^.

Since the curves presented in Figure 7 are so similar that it is difficult to see any difference between them, we computed the absolute error between each curve and the “reference” of *t*_1_ = 82. We present the numerical values in Table 9. Notice that the error is actually below the estimated precision of the approximation.

**Table 9:** Absolute difference between the cumulative distribution given by the stochastic simulations and the reference simulation t_1_ = 82. For each t_1_ we computed the error as 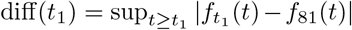, where 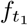 is the estimated distribution computed simulations, for which the initial condition correspond to the components of (2.1) at t = t_1_ rounded to the closest integer.

|  |  |  |  |  |  |  |
| --- | --- | --- | --- | --- | --- | --- |
| $t_1$ | 26 | 33 | 40 | 47 | 54 | 61 |
| date | Jan. 27 | Feb. 3 | Feb. 10 | Feb. 17 | Feb. 24 | Mar. 2 |
| diff($t_1$) | $2.9 \times 10^{-3}$ | $2.1 \times 10^{-3}$ | $2.9 \times 10^{-3}$ | $1.8 \times 10^{-3}$ | $2.5 \times 10^{-3}$ | $1.4 \times 10^{-3}$ |
| $t_1$ | 68 | 75 | 82 | | | |
| date | Mar. 9 | Mar. 16 | Mar. 23 |  |  |  |
| diff($t_1$) | $1.6 \times 10^{-3}$ | $1.2 \times 10^{-3}$ | 0.00 | | | |

## Data Availability

We use the data from WHO

## Author contributions

Q.G., Z.L. and P.M. conceived and designed the study. Q.G. and P.M. analyzed the data, carried out the analysis and performed numerical simulations, Z.L. and P.M. conducted the literature review. All authors participated in writing and reviewing of the manuscript.

## Acknowledgement

The computations presented in this paper were carried out using the PlaFRIM experimental testbed, supported by Inria, CNRS (LABRI and IMB), Université de Bordeaux, Bordeaux INP and Conseil Régional d’Aquitaine (see https://www.plafrim.fr/).

